# Changing species dynamics and species-specific associations observed between *Anopheles* and *Plasmodium* genera in Diebougou health district, southwest Burkina Faso

**DOI:** 10.1101/2024.10.09.24315124

**Authors:** P Lado, LI Gray, E Sougué, AL Knight, M Sorensen, AS Leon, ME Ring, G Pugh, JC Randall, KL Coffin, E Hemming-Schroeder, J Goodwin, M Wade, H Sproch, COW Ouédraogo, SR Dah, AF Somé, RK Dabiré, S Parikh, BD Foy

**Affiliations:** Center for Vector-borne Infectious Diseases, Department of Microbiology, Immunology and Pathology, Colorado State University, Fort Collins, CO, USA; Department of Epidemiology of Microbial Diseases, Yale School of Public Health, New Haven, CT, USA; Center for Innovative Design and Analysis, University of Colorado Anschutz Medical Campus, Aurora, CO, USA; Institut de Recherche en Sciences de la Santé, Direction Régionale de l’Ouest, Bobo-Dioulasso, Burkina Faso

## Abstract

The prevalence of malaria parasite species in parts of Africa is rapidly changing and influenced by detection methods. The natural vector competence and vectorial capacity of African anophelines for human *Plasmodium* species has only been well described for *P. falciparum* and is unclear in the context of mixed and non-falciparum infections. Over the course of two clinical trials (2015 and 2019-2020) testing ivermectin for malaria control in the same region of Burkina Faso, we sampled participants’ blood and their households for *Anopheles* spp. mosquitoes and tested these samples for *Plasmodium* species. *Plasmodium* prevalence in participants and their blood samples was high in both trials. While *P. falciparum* mono-infections comprised most infections in the 1^st^ trial, mixed and non-falciparum infections comprised 27% of infections in the 2^nd^ trial, with notable changes in species present within participants over time. Furthermore, *An. gambiae* s.l. was the main vector captured, but *An. funestus* mosquitoes were unexpectedly prevalent in the 2^nd^ trial, and we found that parasite species prevalence differed in abdominal and head+thorax tissues of these two vector species. Most notably, *P. falciparum* sporozoites were significantly more prevalent than other parasite species in *An. gambiae* s.l. while *P. ovale* sporozoites were significantly more prevalent than other parasite species in *An. funestus*. Our data suggest differential vector competence for *Plasmodium* species at the study site, which may significantly impact malaria epidemiology, disease prevalence and control efforts.

## Introduction

Malaria, caused by *Plasmodium* parasites transmitted by *Anopheles* mosquitoes, is one of the most prevalent infectious diseases in Africa and other parts of the tropics. Of the estimated 249 million cases worldwide, 48.5% occur in West Africa. Burkina Faso is among the most affected countries, ranking 6^th^ in malaria cases and 9^th^ deaths globally (1). Efforts to control the disease burden in the country, similar to other high transmission countries in West Africa, includes using point-of-care (POC) rapid diagnostic tests (RDT) and treatment with artemisinin-based combined therapies (ACT), intermittent preventative treatment in pregnancy (IPTp), and use of seasonal malaria chemoprevention (SMC) with sulfadoxine-pyrimethamine (SP) plus amodiaquine (AQ). To control transmission, health authorities perform community distributions of insecticide-treated bed nets (ITN) approximately every 2-4 years, which more recently includes distribution of nets containing more than one chemical (dual-chemistry nets), and less often conduct indoor residual spraying (IRS) of insecticides in specific regions (2, 3). Despite these efforts, the malaria burden in Burkina Faso has remained high with increasing incidence for nearly a decade (4, 5).

Potential causes for the intransigence of malaria in Burkina Faso despite these intensive control efforts are many. Socioeconomic factors such as high rates of poverty, limited community uptake and/or compliance with parasite and vector control tools, political upheaval with high migrant flows, and extensive artisanal gold mining across the country all likely contribute to the current situation (6–11). Within- and between-species parasite diversity is also underappreciated and represents an additional challenge of malaria control in high burden countries. Infections with a diversity of genotypic clones of *Plasmodium falciparum* (*Pf*) are common in such settings and correlated to higher malaria incidence among young children in many settings (12–14). In center-west Burkina Faso, higher diversity (multiplicity of infection; MOI) of *Pf* genotypes among those presenting with malaria correlated with periods of higher malaria incidence, and overall MOI decreased with advancing age (15). In southwestern Burkina Faso, using amplicon deep sequencing, we recently demonstrated that asymptomatic individuals harbored a mean of 3.1 distinct *Pf* clones per sampling timepoint spanning across wet and dry seasons, each of which could theoretically have different genetics relating to antimalarial drug resistance, immunogenicity, or other factors relating to disease processes (16).

Parasite species diversity also complicates malaria control, but a recognition of the scope of such diversity within different African regions is only recently emerging. Reports over the past several years have demonstrated unexpected or increasing prevalence of *Plasmodium malariae (Pm)*, with its unique 72-hour life cycle, and *P. ovale (Po)* species *and P. vivax (Pv)* with their dormant hypnozoite stages in certain regions (17–19). More limited data from West Africa suggest unexpected prevalence or increases in non-falciparum species in parts of Burkina Faso and other countries (20–22). The lack of detection of non-falciparum species is not surprising because these species tend to maintain lower peripheral parasite densities and therefore are less likely to be detected by microscopy and RDTs (23–25). In addition, African countries nearly universally lack treatment policies which support anti-relapse treatment of dormant hypnozoites of *Po* species and *Pv*, leaving a persistent and unaddressed reservoir within the continent (26).

On the vector side, there is similarly high diversity both within and between mosquito species across Burkina Faso which varies from the dry Sahel ecozone in the north with a short but intense rainy season to the Sudanian ecozone in the south with a longer rainy season (27–37). *Anopheles gambiae* s.s. and *An. coluzzi* typically have the highest human biting rates in rural and semi-rural areas during the rainy seasons (38, 39), while *An. arabiensis* drives malaria transmission in urban centers especially during the dry seasons (32). In other landscapes, different species can be the primary vectors, including *An. funestus* group species around areas with more permanent water sources such as dams, swamps and ponds (39–42), and *An. nili* which breeds along the edges of rivers and streams in the south (43). Highly zoophagic and exophagic vectors such as *An. squamosis*, *An. rufipes*, *An. coustani*, and *An. pharoensis* can also be prevalent across landscapes in the south (43). These diverse vector species and their varied bionomics can hamper vector control efficacy. For example, exophilic and exophagic behaviors that limit exposure to ITN and IRS insecticides, plus increasing insecticide resistance among these vectors due to exposure from both agriculture and malaria vector control efforts, may be hampering the success of the limited vector control tools available (44).. The complexity of parasite transmission from these diverse vectors also likely fosters malaria risk. For example, malaria risk increases with vector density and blood feeding rates among persons with low ITN use (45), and infection intensity and high MOI in different vector species can also drive differential malaria risks or infections with unique genotypes (16, 46–49)

To develop new malaria control tools that might better control malaria in Burkina Faso, we have conducted cluster-randomized control trials in the southwestern Diebougou district to test if ivermectin mass drug administrations (MDA) can reduce malaria incidence. Ivermectin is a widely used endectocide that exhibits mosquitocidal activity (50). The drug’s strong safety profile in humans and broad-spectrum anti-parasitic effects has allowed for its use in MDA to control neglected filarial diseases (51), and ongoing efforts to test its ability to reduce the survival of malaria vectors and limit malaria transmission (52–57).The first trial in Burkina Faso, termed Repeat Ivermectin Mass Drug Administrations for control of Malaria (RIMDAMAL), was conducted from July to November in 2015 and evaluated repeated low dose ivermectin MDA given at 3-week intervals in treatment clusters compared to a single MDA in control clusters (58), while the second trial, termed RIMDAMAL II, occurred in the same region over two consecutive rainy seasons (from July to November in both 2019 and 2020) and evaluated high dose ivermectin or placebo MDA given monthly (59). Additionally, in RIMDAMAL II, MDAs were operationally paired with SMC distribution to children 3-59 months old in all clusters, which was first implemented in the study area in 2018, in between the time of the two RIMDAMAL trials. The RIMDAMAL trials are among several other cluster-randomized control trials that have tested or are in the process of testing if ivermectin MDA can reduce malaria incidence or parasite prevalence among participants in treated communities (60–62).

The outcomes of the RIMDAMAL trial have been published, and the primary and secondary outcomes of the RIMDAMAL II trial will be presented elsewhere. In the process of analyzing the parasite and vector samples collected from these trials, we observed noticeable differences in the prevalence of different parasite species’ infecting participants between the first and second trial, some of whom participated in both trials. Most notably, in RIMDAMAL II, we observed significant associations of specific vector species with different *Plasmodium* species that are suggestive of differences in vector competence and/or vectorial capacity for certain parasite species. In sub-Saharan Africa (SSA), despite *Pf* being the species most frequently detected and responsible for the majority of clinical cases (63), *Po* and *Pm* can significantly contribute to malaria epidemiology and have been observed to occur at high prevalence in parts of several countries (18, 19, 21, 64–73) or increasing in prevalence over longitudinal studies within a country (20). The data presented here shed light on vector-parasite interactions that may influence malaria epidemiology observations in these and other studies.

## Results

### Organization and timelines surrounding the RIMDAMAL trials

The health district of Diebougou is in the southwest region of Burkina Faso in Bougouriba province. The district has been conducting yearly MDA with single dose ivermectin (150 µg/kg) and albendazole (400 mg/kg) in all its communities for more than a decade to control onchocerciasis and lymphatic filariasis. The RIMDAMAL trial was conducted during the 2015 rainy season in 8 clusters (villages) in the health district, while the RIMDAMAL II trial was conducted during the 2019 and 2020 rainy seasons in 14 clusters of the district. Trial designs, cluster selection and allocation in both trials are described elsewhere (58, 59). Briefly, in both trials, clusters were randomized 1:1 between the treatment and control arms, and the trial organizations and treatments and controls were different, however four clusters were common among the two trials (Table 1). In RIMDAMAL, repeated low dose ivermectin MDA (single oral dose, 150 µg/kg) was given at 3-week intervals in treatment clusters and compared to a single MDA at the start of the trial in control clusters, while the RIMDAMAL II trial occurred over two consecutive rainy seasons and repeated high dose ivermectin MDA (1 oral dose daily for 3 days; 300 µg/kg) was given at 1-month intervals in treatment clusters compared to placebo MDA on the same schedule in control clusters. Furthermore, in RIMDAMAL II, MDA was operationally paired with SMC distribution to children between 3-59 months old in all clusters, which was instituted as national policy starting in 2017. A third, unrelated, cluster randomized trial called the REACT trial was conducted from 2016-18 by a few of the authors, which had one trial arm including ivermectin treatment of cattle, goats and sheep in the 2017 rainy season and occurred in villages of the Diebougou health district that were not part of the RIMDAMAL II trial (74, 75). District ITN distributions occurred in 2016 with standard pyrethroid-based nets, and at the end of 2019 (between October 29-November 2) with new dual-chemistry (alpha-cypermethrin and chlorfenapyr) Interceptor^®^ G2 nets to better control insecticide-resistant malaria vectors. An analysis of Normalized Difference Moisture Index (NDMI), a measure of vegetation moisture content and indicator of land surface wetness, across the study areas from 2015-2020 revealed marked interannual variation as well as variation between clusters (Figure 1). Overall, the 2015 RIMDAMAL trial was conducted during a drier rainy season of those analyzed for the study area, while both rainy seasons of RIMDAMAL II (2019 and 2020) were relatively wet (Figure 1). NDMI was selected for analysis because it provides information on both temporal precipitation and landscape heterogeneity which may correspond to differences in larval habitat abundance (76).

**Table 1.**
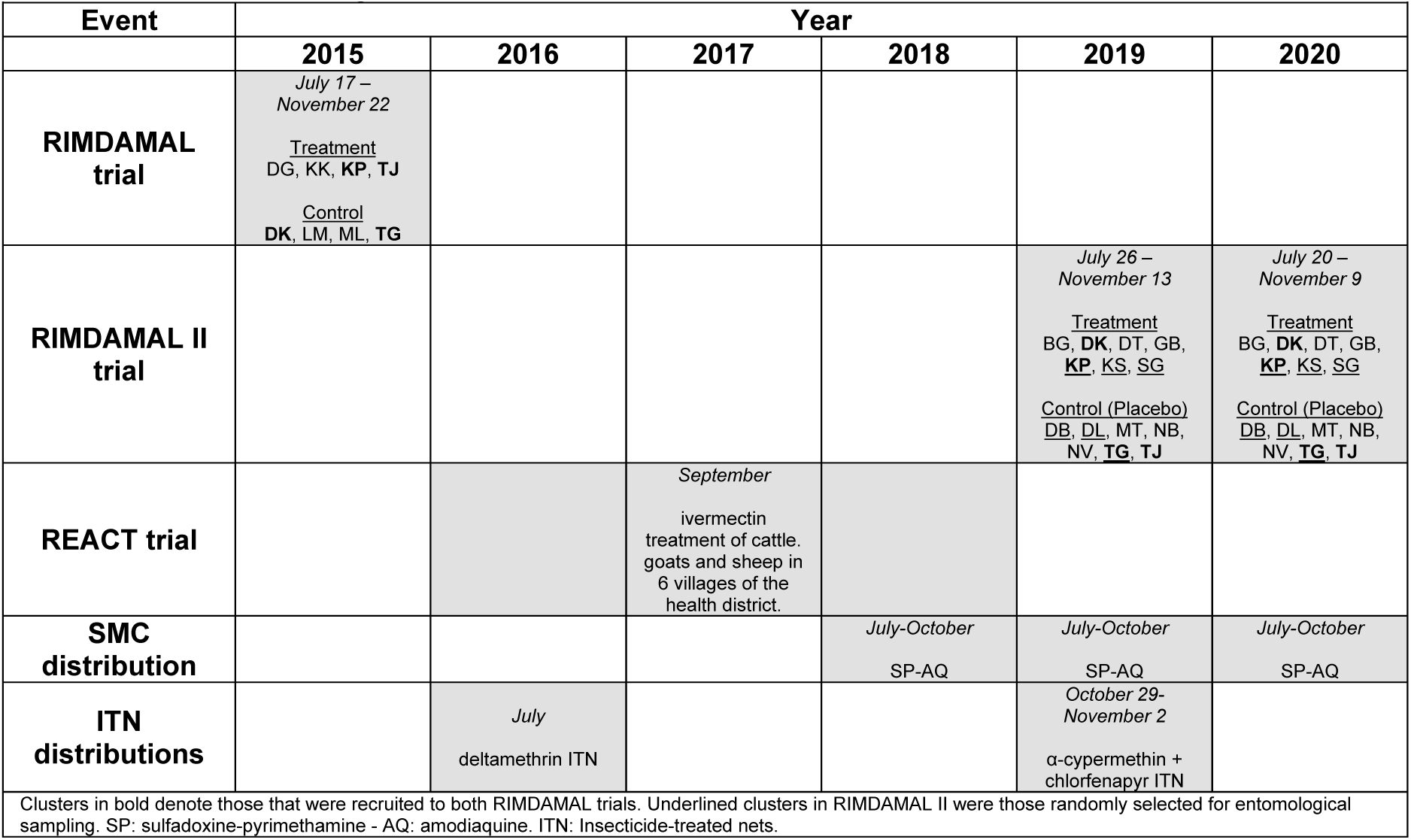
Timeline and organizational features of the RIMDAMAL trials and other malaria control trial and efforts in the Diebougou health district.

**Figure 1.**
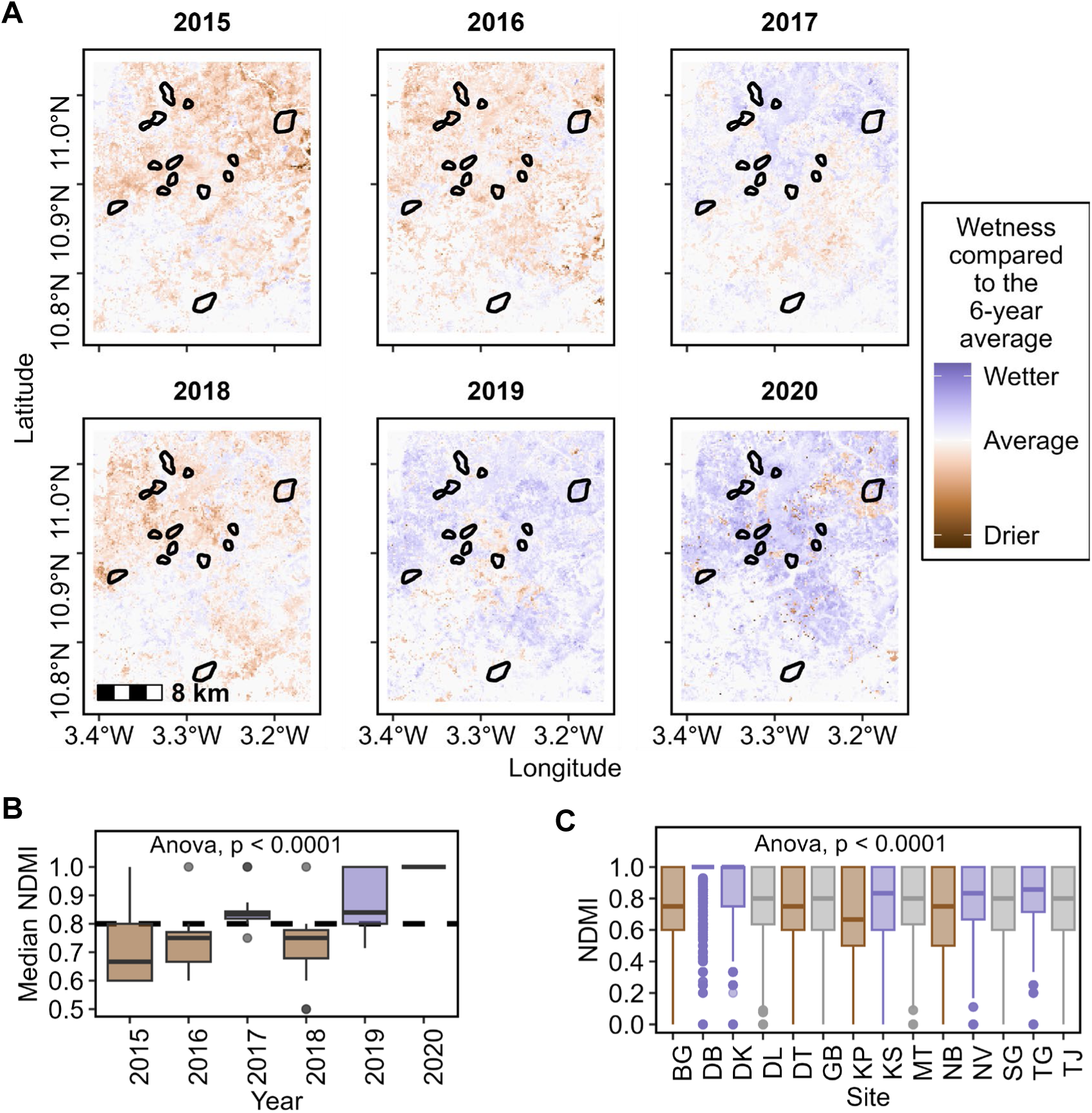
Spatiotemporal variation in wetness inferred from Landsat-8 Normalized Difference Moisture Index (NDMI). Satellite images (Landsat 8 Level 2, Collection 2, Tier 1) were obtained from the rainy seasons (May to Oct) and median NDMI calculated for each year. **A)** Map of relative wetness at the study site and clusters (outlined in black) **B)** Temporal variation in mean study site NDMI. Dashed line indicates overall median value. **C)** Spatial variation in NDMI at each cluster (all years).

### Differences in the prevalence of *Plasmodium* species between participants of the RIMDAMAL and RIMDAMAL II trials

The prevalence of blood smear slides from the 2015 RIMDAMAL trial that were positive by microscopy for *Plasmodium* spp. and for different *Plasmodium* species was previously reported, as was the multiplicity of infection and molecular force of infection in *Pf* positives (58). Here we report data using molecular analyses on 90 archived blood samples taken from 24 children aged 0-5 years (split equally from 2 clusters in each arm) who had a fever or history of fever in the prior 24 hours. Of those, 39% (35/90) of tested samples were positive for *Plasmodium* species and all were *Pf* mono-infections. Overall, 71% (17/24) of these tested children had a molecularly confirmed parasitemia at least once during the trial (Table 2).

**Table 2.** Molecular detection of *Plasmodium* parasites from participants of the RIMDAMAL trials.

|  |  |  |  |  |  |  |  |  |  |  |  |  |  |  |  |  |
| --- | --- | --- | --- | --- | --- | --- | --- | --- | --- | --- | --- | --- | --- | --- | --- | --- |
| Trial | Participants (any longitudinal sample) |  |  |  |  |  |  |  | Participants' samples |  |  |  |  |  |  |  |
|  | ACD cohort (0-5 yrs old)* |  |  |  | Participants ≥ 6 yrs old |  |  |  | ACD cohort (0-5 yrs old)* |  |  |  | Participants ≥ 6 yrs old |  |  |  |
|  | *Pf* | *Pf*+*nPf* | *nPf* | neg | *Pf* | *Pf*+*nPf* | *nPf* | neg | *Pf* | *Pf*+*nPf* | *nPf* | neg | *Pf* | *Pf*+*nPf* | *nPf* | neg |
| RIMDAMAL | 71%<br>(17/24) | 0%<br>(0/24) | 0%<br>(0/24) | 29%<br>(7/24) | - | - | - | - | 39%<br>(35/90) | 0%<br>(0/90) | 0%<br>(0/90) | 61%<br>(55/90) | - | - | - | - |
| RIMDAMAL II | 59%<br>(16/27) | 22%<br>(6/27) | 11%<br>(3/27) | 7%<br>(2/27) | 37%<br>(16/43) | 53%<br>(23/43) | 2%<br>(1/43) | 7%<br>(3/43) | 30%<br>(47/156) | 5%<br>(8/156) | 4%<br>(6/156) | 61%<br>(95/156) | 46%<br>(114/249) | 16%<br>(40/249) | 4%<br>(10/249) | 34%<br>(85/249) |
| *ACD: active case detection cohort. In RIMDAMAL, SMC was not yet initiated, but in RIMDAMAL II eligible members of the ACD cohort (those 3-59 month old) were given SMC. *Pf*: *P. falciparum*, *nPf*: *P. malariae*, *P. ovale*, *P. vivax*. |  |  |  |  |  |  |  |  |  |  |  |  |  |  |  |  |

\*ACD: active case detection cohort. In RIMDAMAL, SMC was not yet initiated, but in RIMDAMAL II eligible members of the ACD cohort (those 3-59 month old) were given SMC. *Pf*: *P. falciparum*, *nPf*: *P. malariae*, *P. ovale*, *P. vivax*.

In contrast, in the RIMDAMAL II trial, infections with multiple different *Plasmodium* species were commonly detected in human dried blood spots from 70 randomly chosen RIMDAMAL II participants (split equally from 3 clusters in each arm, and between age grouping and sex) from our cross-sectional cohort (3 villages per arm) sampled over 2 seasons (2019–2020) (Table 2). Of the 405 samples tested, 56% (225/405) were positive for *Plasmodium* species, and of these 72% (161/225) were *Pf* mono-infections and 28% (64/225) were *Pf*-mixed or non-falciparum infections (12% (27/225) *Pf+Pm* co-infections, 5% (12/225) *Pm* mono-infections, 5% (12/225) *Pf + Po* co-infections, 2% (4/225) *Po* mono-infections, 0.4% (1/225) *Pf + Pv* co-infections, and 3% (7/225) *Pf+Po+Pm* infections). Children 0-5 years old (who took SMC in the trial) more often had negative blood samples over the trial period relative to participants ≥ 6 years old (61% vs. 34%; P < 0.0001) and tended to less often be infected with at least one non-falciparum species over the trial period relative to participants ≥ 6 years old (56% vs. 33%; P = 0.0871). Across all age groups, most non-falciparum infections were detected in one or two cross-sectional blood samples obtained from individuals throughout the trial period (Figure 2).

**Figure 2.**
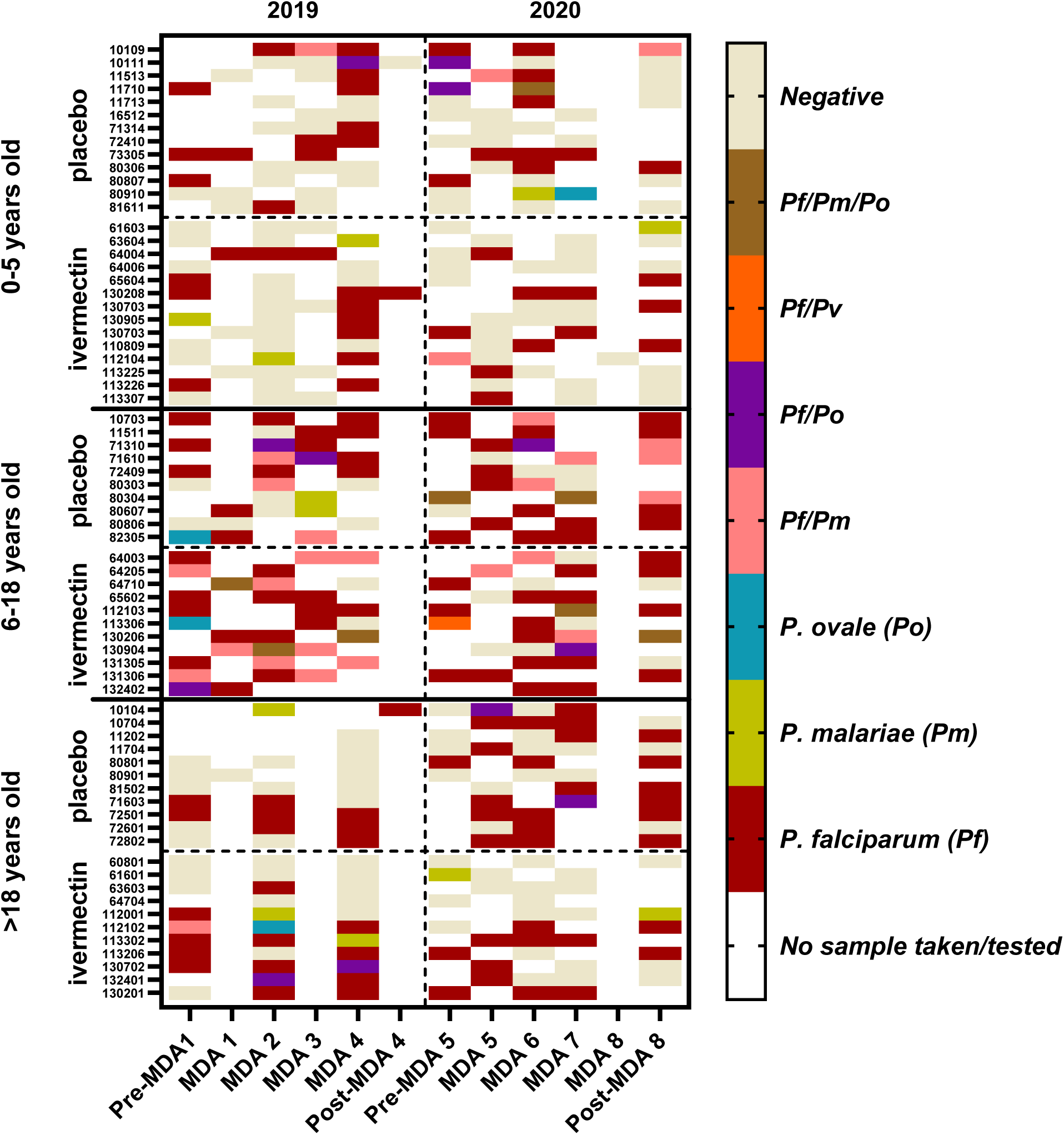
Parasite species detected in human dried blood spots from participants of the RIMDAMAL II trial. De-identified codes of participants are shown on the Y-axis. Participants from the 6 clusters (3 ivermectin-treated and 3 placebo-treated) that were randomly selected for entomological and cross-sectional blood sampling during the trial were selected from those who had DBS sampled across the entire study period, and who were equally dispersed between three age groups and genders. Time points on the X-axis refer to the 28-day period in between MDA (mass drug administrations) or approximately 1 week pre- or post-the MDA intervention periods per season.

### *Anopheles* speciation and blood meal analysis

RIMDAMAL entomology data were previously reported (58). Briefly, 2,801 mosquitoes were captured via indoor aspirated of which 99% (2,784/2,801) were morphologically identified as *An. gambiae* s.l. These were blood fed mosquitoes that were separated into 1) head+thorax tissue that were previously homogenized and used in sporozoite ELISA assays to detect *Pf* circumsporozoite (CSP) protein, and into 2) bloodfed abdomens that were squashed onto filter papers cards but then used in other experiments.

In RIMDAMAL II, a total of 7,942 anopheline mosquitoes were collected (2019, n = 5,827; 2020, n = 2,115) of which the majority were indoor-captured blood-feds that were held in assays to measure their survival over time. Morphological identification determined the majority to be *An. gambiae* sensu lato (s.l.) (85%; 6,745/7,942), with approximately twice the number captured in 2019 (n = 4,679) as compared to 2020 (n = 2,066). The remainder were *An. funestus* group mosquitoes (15%; 1,197/7,942) of which 96% (1,148/1,197) came from 2019. A subset of *An. gambiae* s.l. mosquitoes were further speciated using molecular methods (n= 354). The majority were identified as *An. gambiae* (79.4%), followed by *An. coluzzi* (15.5%) and *An. arabiensis* (5.1%). Similarly, a subset of *An. funestus* group mosquitoes were further speciated (n = 251) and the majority were identified as *An. funestus* (96.4%), while a small portion were identified as *An. rivulorum* (3.2%) and *An. leesoni* (0.4%).

Blood meal analysis was performed on bloodfed mosquito abdomens from a subset of *An. gambiae* s.l. and *An. funestus* complex mosquitoes (n = 171 and 64, respectively). Human blood meals were detected in most samples from both *An. gambiae* s.l. (81%) and *An. funestus* (68%) (P = 0.0559). Cattle blood meals were the next most abundant but were significantly more prevalent in *An. funestus* (23%; 15/64) than in *An. gambiae* s.l. (6%; 11/171) (P = 0.0006). Dog, goat, and pig blood meals were detected in both mosquito species, but were uncommon (Figure 3).

**Figure 3.**
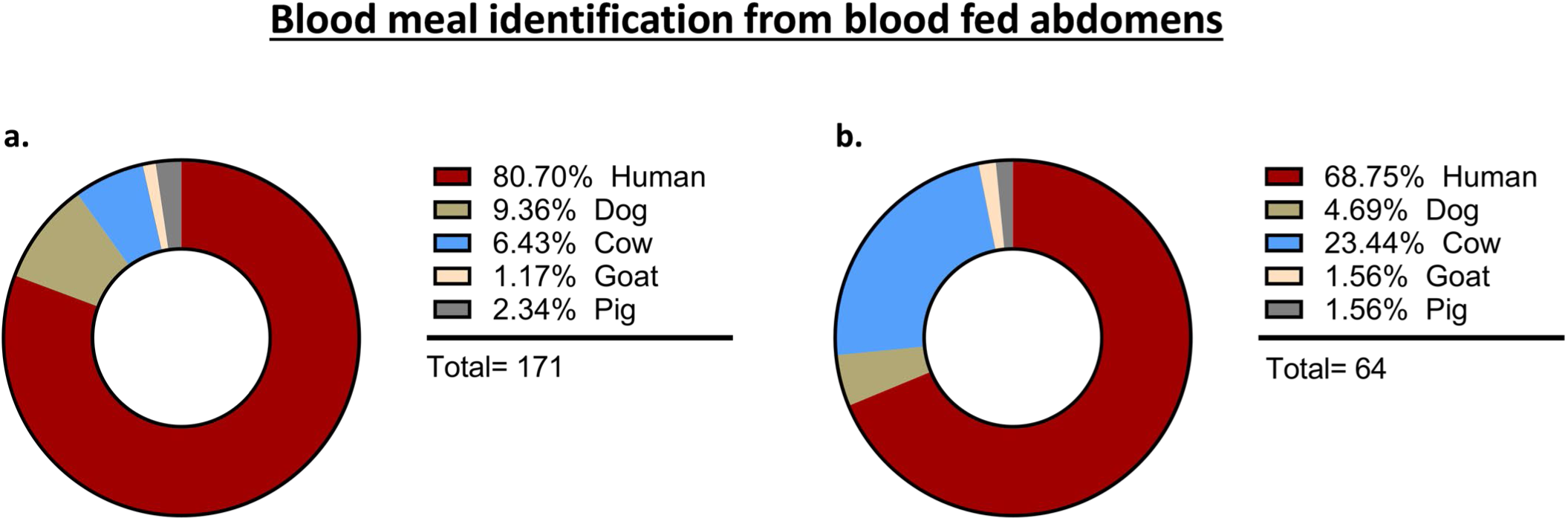
Blood meal identification from blood fed mosquitoes captured in participant households sampled in the RIMDAMAL II trial. a) *Anopheles gambiae* s.l and b) *Anopheles funestus*.

### Prevalence of *Plasmodium* parasites species detected in bloodfed and unfed mosquito head-thorax tissues from both trials

In RIMDAMAL, the overall sporozoite rate over the intervention period and across arms was 4.8% (62/1759; *An. gambiae* s.l. only, head-thoraxes tested by CSP-ELISA) (Table 3).

**Table 3.** Sporozoite rates in the RIMDAMAL trials.

| Table 3. Sporozoite rates in the RIMDAMAL trials. |  |  |  |  |
| --- | --- | --- | --- | --- |
| Trial | Mosquito species | Trial arm | Sporozoite rate over trial | P value |
| RIMDAMAL | *An. gambiae* s.l. | Control | 3.4% (33/961) | - |
|  |  | Treatment | 3.6% (29/798) | 0.8207 |
| RIMDAMAL II | *An. gambiae* s.l. | Control | 5.3% (74/1399) | - |
|  |  | Treatment | 4.3% (48/1129) | 0.2626 |
|  | *An. funestus* | Control | 20.4% (23/113) | - |
|  |  | Treatment | 11% (15/136) | 0.0416 |
|  | *An. gambiae* s.l. +<br>*An. funestus* | Control | 6.4% (97/1512) | - |
|  |  | Treatment | 5% (63/1265) | 0.1201 |
| Sporozoite detection from RIMDAMAL trial was performed using head-thoraxies in CSP-ELISA assays using a monoclonal antibody against *Pf*, while sporozoite detection from RIMDAMAL II was performed using head-thoraxies in qPCR assays designed to detect *Pf*, *Po*, *Pm* or *Pv* species. |  |  |  |  |

From RIMDAMAL II, a total of n = 2,777 mosquito head-thorax samples (n = 2,528 *An. gambiae* s.l and n = 249 *An. funestus*) were screened for *Plasmodium* sporozoites using species-specific qPCR. The overall sporozoite rate over the intervention period and across arms was 5.8 % (n = 160/2,777). In *An. gambiae* s.l. the overall sporozoite rate across arms was 4.8 % (n = 122/2528) and the most common *Plasmodium* species of sporozoites detected among positives was *Pf* mono-infections (65%; 79/122), followed by *Po* mono-infections (25%; 31/122), *Pm* mono-infections (8%; 10/122), and only two mixed species infections (both *Pf+Po* co-infections; Figure 4c). In contrast, *An. funestus* had a significantly higher overall sporozoite rate over the intervention period than *An. gambiae* s.l. (15.3%; 38/249; P < 0.0001). Also, in contrast to *An. gambiae* s.l., the most prevalent sporozoite species detected among positives of *An. funestus* was *Po* (63.2%;24/38), followed by *Pf* and *Pm*, both at 18.4% (7/38 each) (Figure 4f). Thus, in the RIMDAMAL II trial, the *Pf* sporozoite rate in *An. gambiae s.l.* was significantly greater than in *An. funestus* (P <0.0001), and the *Po* sporozoite rate in *An. funestus* was significantly greater than in *An. gambiae s.l.* (P <0.0001) from mosquitoes captured in the same villages and houses over the same dates. Furthermore, while the overall sporozoite rates and those from *An. gambiae s.l.* captured in clusters from treatment or control arms were not significantly different from one another, sporozoite rates were significantly higher (∼2 fold) in the control arm relative to the intervention arm for *An. funestus* (P = 0.0416; Table 3).

**Figure 4.**
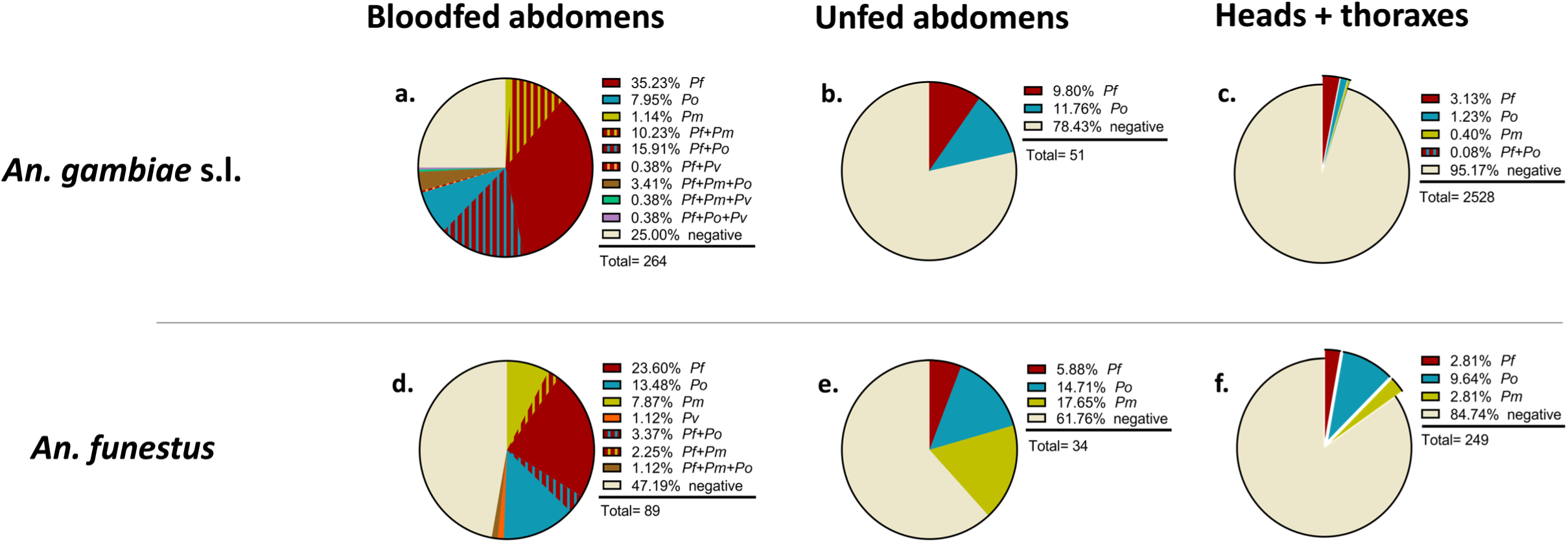
Parasite species detected in different tissues of *Anopheles gambiae* s.l and *Anopheles funestus* collected from households of sampled clusters in the RIMDAMAL II trial. Mosquito tissues tested were dissected blood fed abdomens (a & d) containing fresh blood and primarily caught using indoor aspirations, dissected unfed abdomens (b & e) captured using both indoor aspirations as well as light traps placed indoors and outdoors, and head-thoraxes (c & f) captured using indoor aspirations as well as light traps placed indoors and outdoors.

### Prevalence of *Plasmodium* parasites species detected in bloodfed and unfed mosquito abdomens in RIMDAMAL II

To better understand the observed differences in sporozoite rates between species and treatment arms in RIMDAMAL II, we examined a portion of the stored bloodfed and unfed abdomen samples from each mosquito species and from those that were either sporozoite-positive or sporozoite-negative. Of the 264 *An. gambiae* s.l. bloodfed abdomens tested, 75% (198/264) were positive for at least one *Plasmodium* species (Figure 4a). Approximately half of positives (47%; 93/198) contained parasites of only one species and most often this was *Pf*. However, nearly as many (41%; 81/198) contained mixed parasites consisting of *Pf* plus one or more non-falciparum species. In contrast, of the *An. funestus* bloodfed abdomens tested, a significantly lower proportion were *Plasmodium*-positive (52.8%; 47/89; P = 0.0002) (Figure 4d), with the majority (87.2%; 41/47; P < 0.0001) containing parasites of a single species of either *Pf*, *Po* or *Pm.* Overall *Plasmodium* spp. prevalence in blood fed abdomens was modestly higher in sporozoite-positive versus sporozoite-negative mosquitoes, regardless of mosquito species (Figure 5). Similarly, mosquitoes captured from treatment arm clusters had a modestly lower prevalence of *Plasmodium* spp. than those captured from control arm clusters (Figure 6).

**Figure 5.**
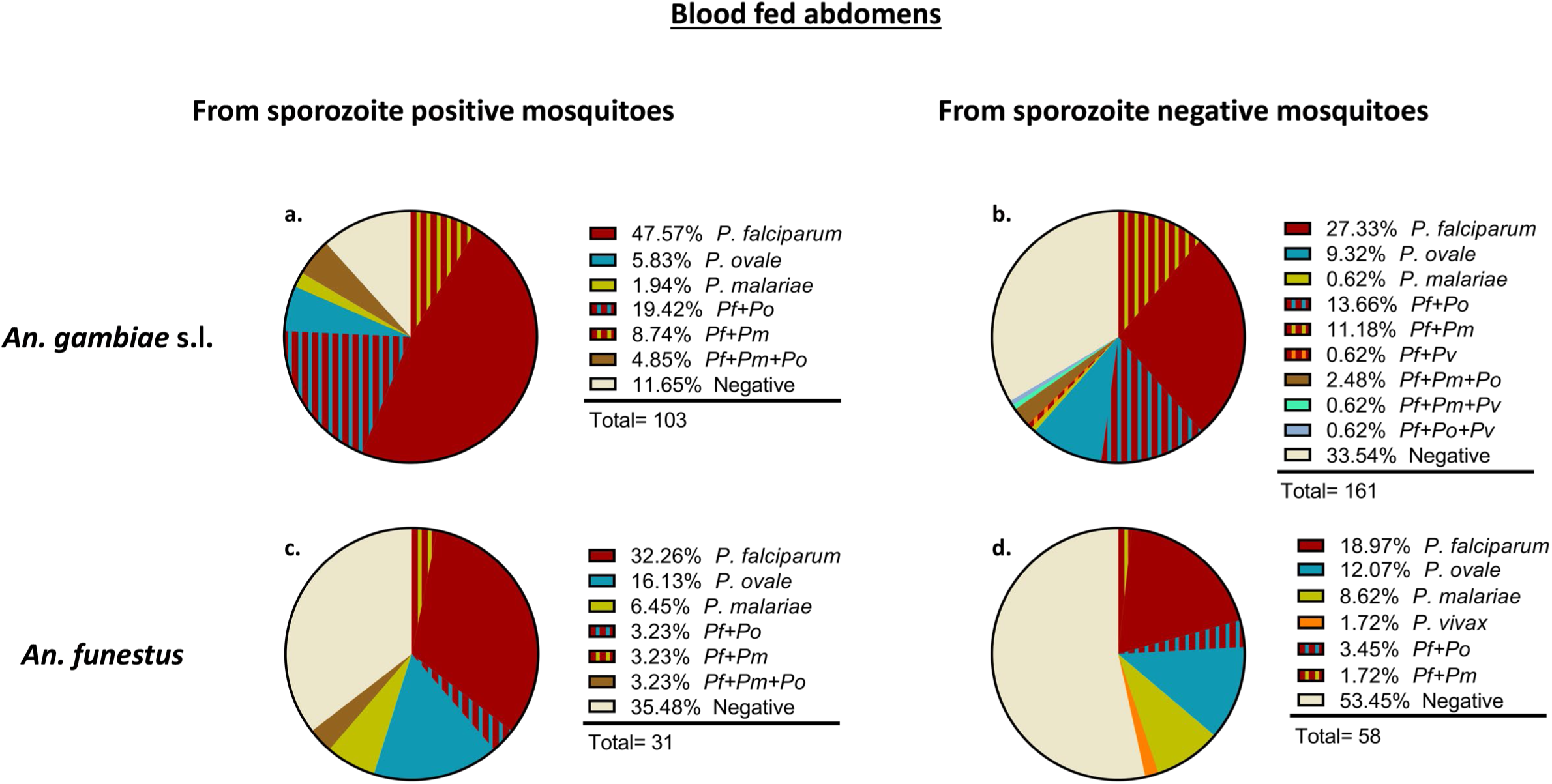
Parasite species detected in the blood fed abdomens of mosquitoes collected from households of sampled clusters in the RIMDAMAL II trial. The blood fed abdomens are from either sporozoite positive (a & c) or sporozoite negative (b & d) *Anopheles gambiae* s.l (a & b) and *Anopheles funestus* (c & d).

**Figure 6.**
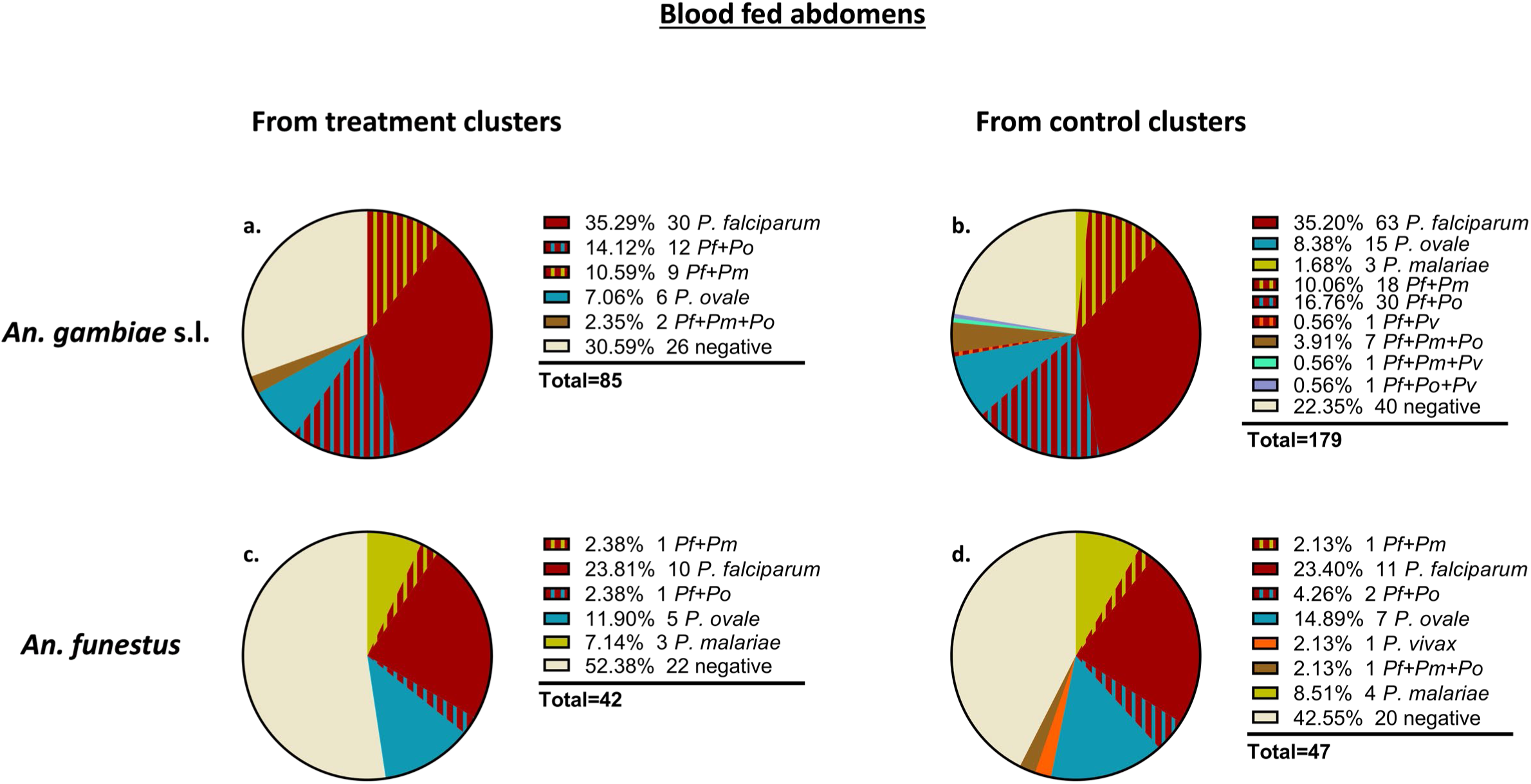
Parasite species detected in the blood fed abdomens of mosquitoes collected from households of sampled clusters from the different arms the RIMDAMAL II trial. The blood fed abdomens were mosquitoes caught from either treatment (ivermectin) clusters (a & c) or control (placebo) clusters (b & d) and were from *Anopheles gambiae* s.l (a & b) and *Anopheles funestus* (c & d).

Most unfed abdomens tested negative for parasites (Figure 4b & 4e), in contrast to the blood fed abdomens of the same species (Figure 4a & 4c). Furthermore, *Plasmodium* species prevalence in these unfed abdomens were higher than, but reflective of, the sporozoite-positive head-thoraxes of the different mosquitoes in that only mono-infections were detected. *Pf* and *Po* infections were approximately equal among the unfed abdomens of these mosquitoes, but *Pm* infections of unfed abdomens were only detected in *An. funestus* (Figure 4b & 4e).

## Discussion

The results presented suggest that malaria epidemiology at the study site appears to have changed over a six-year period whereby non-falciparum species have become prevalent, particularly as mixed infections with *Pf*. The data also suggest that vectors species have differential competence for spreading non-falciparum infections, which may be one novel driver explaining changes in malaria species epidemiology. Unfortunately, standard malaria surveillance performed by the national health authorities, and most other malaria studies in the country over the last decade, have focused on tackling the burden of *Pf,* and molecular data on the prevalence of non-falciparum species in Burkina Faso is largely lacking. One other published study, however, similarly reported an increase in non-falciparum infections from 2007-2010 in central-west Burkina Faso in, but entomological indices were not reported (20).

In addition to entomological factors, anthropogenic factors regarding diagnostics and treatment algorithms have likely driven or influenced the epidemiologic changes observed. Most notably, current POC diagnostics suffer from relatively low sensitivity, requiring peripheral densities of ∼100-500 parasites/μL for detection, yet most non-falciparum species tend to maintain lower parasitemias that go undetected (17, 18, 23, 24, 77–79). In addition, while microscopy has the advantage of morphologic-based speciation, this is challenging in practice, particularly in the presence of mixed species infection. With regards to RDTs, nearly 80% of RDTs currently procured in sub-Saharan Africa detect only *Pf-*specific HRP-2 (80–82). While a second less widely available RDT detects both *Pf*HRP-2 and pan-*Plasmodium* LDH (83–86), its sensitivity is also low at densities below 1000/μL (25, 87–89).Treatment and prevention guidelines also likely impact species prevalence, as most strategies are explicitly geared towards reducing the *Pf* burden. In West Africa, the implementation of SMC, including at the site of the RIMDAMAL trials in 2018, has proven to have significant impacts on *Pf* incidence in children, and although it is anticipated that SP-AQ has activity against non-falciparum species, its precise impact on these other species has not been investigated (90–96). Similarly, the impact of ACTs on non-falciparum species is less well studied, and the near-complete lack of usage of 8-aminoquinolines directed at the dormant liver stages of *Po* and *Pv* in Africa results in a persisting reservoir of infection even if blood-stage therapy is initiated. In a 2-year study of malaria prevalence in Niger during and after SMC distributions, cross sectional microscopy surveys detected *Pf* alone in the high and low seasons of year 1, but subsequently detected mixed- and mono-infections with *Po* and *Pm* in year 2 (97). In our RIMDAMAL trials, incidence in 0 to 5 year olds dropped approximately 80% between 2015 and 2019 (58) (ClinicalTrials.gov: NCT03967054), presumably due to SMC commencing in 2018, but parasite prevalence remained exceedingly high in both trials suggesting a limited ability of SMC to affect parasite prevalence in this high transmission setting. Malaria incidence among the child cohort in RIMDAMAL II dropped further in 2020 across both arms, which is likely connected to the activity of dual chemistry nets distributed in both trial arms. It is also possibly that an increase in non-*falciparum* parasite species at the field site may have contributed to the overall reduction in malaria incidence via our active case detection procedures between the two RIMDAMAL trials given that the pyrogenic thresholds of non-*falciparum* species are likely to be lower than *Pf* (98) from considerably lower parasitemias and shorter durations of patent infections (23, 24, 99, 100).

The abundance of *An. funestus* in RIMDAMAL II during the 2019 rainy season was unexpected considering our data from previous seasons (39, 41, 58). Typically, this vector increases at the very end of the rainy season and persists through the early, ‘cold’ dry season (late October - February), a time when *An. gambiae* s.l. populations fall dramatically. Its presence is connected to more permanent water sources with emergent vegetation (42, 101) that remain following the cessation of the seasonal rains. *An. funestus* was highly prevalent at the study site in human landing catches performed during dry season sampling between 2017-8 during the REACT trial (75). Its abnormally high density so early in the year in 2019 may be related to the wetter-than-average rainy season (Figure 1) where it could take advantage of the more abundant, possibly deeper water breeding sites across the landscape due to the excess water. Regardless, the opportunity to catch both *An. gambiae* s.l. and *An. funestus*, especially the blood feds, in the same households over the same timeframes allowed for novel insights into potential differential vector competence between *An. gambiae* and *An. funestus* in our area.

The tested blood fed abdomens of both mosquito species would be expected to contain a mix of newly ingested parasites (both asexual forms and gametocytes) from mostly human blood meals that were taken the night prior to capture, as well a smaller proportion of developing early sporogonic stages in and on the midgut tissues of mosquitoes that were infected from an earlier blood meal. The unfed abdomens, being from host-seeking mosquitoes captured in the same households as the blood feds, provided a measure of the proportion of mosquitoes already containing those developing sporogonic stages, which was 21.6% for *An. gambiae* s.l. and 38.2% for *An. funestus*. Considering the proportions of negative abdominal tissues between blood feds and unfeds, the data suggest *An. gambiae* s.l. are exposed to parasites upon blood feeding nearly twice as often as *An. funestus*, likely from higher human biting rates that we observed from this mosquito, and potentially also from a higher propensity to blood feed on multiple people over a single night (102). The data also show that mixed-parasite species blood meals, which we surprisingly observed often in the human samples too, were present nearly five times as often among *An. gambiae* s.l. than *An. funestus*, but mixed infections were not detected in unfed abdomens of either mosquito species, and only detected twice in thoraxes as sporozoites. Dividing the unfed abdomen infection rates by the blood fed abdomen parasite rate can provide a rough approximation of successful midgut infection in the wild for both mosquito species, which was only 29% of the time for *An. gambiae* s.l. but approximately 72% of the time for *An. funestus*. Examination of infected unfed abdomens compared to sporozoite infections in the positive head+thorax tissues suggests that successful parasite development from early to late sporogonic forms may be similarly more restrictive in *An. gambiae* s.l. (22%) than in *An. funestus* (40%). Lastly, the differences observed in the prevalence of different *Plasmodium* species across the tissues of the two mosquitoes is notable. These data suggest that while *An. funestus* was less often exposed to parasites during blood feeding, the exposure was more often as mono-infections and parasite infection was more often successful. Furthermore, *An. funestus* was preferentially infected with *Pm* or *Po*, but complete sporogonic development to sporozoites more often favored *Po*. In contrast, *An. gambiae* s.l. captured at the same timepoints and from the same households was more often exposed to parasites during blood feeding and up to 40% may have been mixed-infection exposures, but parasite infections were less successful. Furthermore, midgut infections in *An. gambiae* s.l. (unfed abdomens) were nearly always of a single parasite species which was typically *Pf* and *Po,* but *Pf* more often successfully developed into sporozoites.

Given these are observational entomological and human data, there are several non-mutually exclusive hypotheses that could explain the findings. Most obviously, data may reflect true vector competence differences of mosquito species for different parasite species, such as parasite exflagellation, fertilization or sporogonic development being enhanced for *Po* in *An. funestus*, or the inverse whereby these are inhibited for *Pm* and *Po* in *An. gambiae*, perhaps via innate immune targeting mechanisms (103). This may be an observation of emergent co-speciation between vector and parasite species. Parasite species sporogony competition, either directly or for resources in the mosquito, may also contributing to preferential vector competence outcomes in one mosquito species over the other (104–108). Lastly, factors influencing vectorial capacity may also help explain some of these observations. The *An. funestus* we captured blood fed more often on cattle, which may explain some of the parasite prevalence data in blood fed abdomens, but another possibility is that *An. funestus* preferentially blood fed on human hosts with different parasite species relative to *An. gambiae*, possibly due to a preference for certain attractive odors given off by the infected host dependent on the parasite species they were infected with (109). Some of the results may also be explained by differences in vector blood meal sizes or vector lifespans, such as if most of the *An. funestus* mosquitoes are longer-lived and therefore more likely to have non-falciparum sporozoites which develop less rapidly than *Pf*, or mortality in the field due to certain combinations of parasite and vector, for example if *Pm* sporogonic development increases *An. gambiae* s.l. mortality. We are not aware of previous field reports on co-speciation or preferential sporogonic development of certain *Plasmodium species* over others in different species of vector anophelines. Molecular detection of *Plasmodium* in mosquitoes captured from the field in African settings remains relatively rare due to the costs of performing qPCR on numerous individual mosquito samples with different molecular probes. More often ELISA assays are used to save costs, but these are often only used with antibodies to *Pf* or *Pv* alone (110) and are relatively insensitive and prone to false positives when compared to molecular analyses (111–113). Also, for molecular analyses of mosquitoes, the most efficient molecular targets are also still being compared (114, 115), as are the dissection methods to ensure the fewest false positives (116). We chose to dissect at the junction of the midgut and abdomen, however, the large differences in parasite prevalence between blood fed abdomens compared to unfed abdomens and head+thoracies suggests our analyses was not overly biased from parasite contamination across mosquito tissues. Regardless, this was a comparative analysis between mosquito species.

Importantly, this study has several limitations. Foremost, these longitudinal data are confined to a single field site and must be tested to determine their generalizability. It is possible that the results are partially related to the ivermectin or other intervention activities occurring at the site. Regarding the human data, we had limitations from our more limited testing of archived samples from the RIMDAMAL trial, including that the blood samples tested were only from symptomatic children. Retrospective molecular analysis of mosquito tissues from the RIMDAMAL trial were also not possible due to the samples having been used in prior assays. Follow-up blood sampling and testing should be performed at the site to determine if the change in prevalence from *Pf* to more non-*falciparum* species persists or increases over time, and if non-*falciparum* infections are correlated with presence of *An. funestus* or increased rainfall during the wet season in other areas. While the entomological data are a measure of natural infection outcomes, the analyses are associative being based on observational data alone and so they should be validated using mosquito feeding assays with the blood from infectious participants.

## Conclusion

The data presented here show that malaria parasite species prevalence in humans changed dramatically in the Diebougou health district in southwest Burkina Faso over the period bracketing the two RIMDAMAL trials occurring five years apart, and when SMC was initiated at the site in between the two trials. The parasite species changed from almost exclusively being *Pf* to a high prevalence of non-falciparum species mixed with *Pf* or occurring alone.

Furthermore, evidence suggests that non-falciparum species were preferentially transmitted by *An. funestus* while *Pf* was preferentially transmitted by *An. gambiae* s.l. These formative studies should be followed up with more rigorous testing of the hypotheses regarding vector competence or vectorial capacity differences for *Plasmodium* parasites between the different mosquito species and may explain the changing malaria epidemiology in areas of sub-Saharan Africa where a diversity of anopheles vectors exist or are in flux.

## Methods

### Study area, experimental design, and adult mosquito collection

The Diebougou health district in southwestern Burkina Faso experiences a single long rainy season from approximately June-November (39). The RIMDAMAL trial spanned one rainy season (July-November) in 2015, and RIMDAMAL II spanned rainy seasons (July-November) in two consecutive years, 2019 and 2020 and both were conducted in this health district. Detailed information about both trials is available elsewhere (58, 59). Briefly, in RIMDAMAL, 8 village clusters were studied and 4 were randomized to the intervention arm consisting of 6 ivermectin MDAs (single dose of 150 µg/kg) for eligible participants every 3 weeks over one season, while the other 4 were randomized to the control arm consisting of a single ivermectin MDA (single dose of 150 µg/kg) for eligible participants at the beginning of the season. In RIMDAMAL II, 14 village clusters were studied and 7 were randomized to the intervention arm consisting of 4 ivermectin MDA (3 consecutive daily doses of 300 µg/kg) for eligible participants once per month over 4 months of the season, while the other 7 were randomized to the control arm consisting of placebo administered in the same manner. To estimate spatiotemporal variations in land surface wetness in the study area, Normalized Difference Moisture Index (NDMI), a measure of vegetation moisture content, was assessed from 2015-2020. NDMI was measured from Landsat 8 Level 2, Collection 2, Tier 1 satellite images at 30-m resolution. Satellite images were obtained from the rainy seasons (May to October) and median NDMI calculated for each year from near infrared and short-wave infrared bands after cloud masking in Google Earth Engine.

*Anopheline* mosquito sampling in both trials was performed similarly, in 8 households from each village in RIMDAMAL and from 8 households in each of 3 villages per arm in RIMDAMAL II, whereby most mosquitoes captured were resting indoor blood feds and aspirated from walls and ceilings in the mornings, and additional capture of host-seeking mosquitoes was performed using light traps located a single centrally located house per village placed indoors next to a person sleeping under a bed net and outdoors next to a person sleeping in a tent. In RIMDAMAL, mosquito collections occurred on the 2^nd^ week follow each MDA, and in RIMDAMAL II on the 1^st^ and 3^rd^ weeks following each MDA. Participants’ blood from these same households was also serially sampled across the intervention periods to allow linking of parasitology and entomology outcomes. After collection, mosquitoes were morphologically identified in the field station as either *An. gambiae* s.l., *An. funestus* complex, or other species, and molecular analyses were performed subsequently in laboratories.

### DNA extraction from dried human blood spots

Dried blood spots (DBS) on filter paper cards were made in RIMDAMAL from active case detection (ACD) cohort children whose blood was sampled during febrile episodes occurring during the trial. DBS from 24 of these randomly chosen participants who were living in the 4 villages that were also enrolled in RIMDAMAL II (Table 1; KP, TJ, DK, TG) and which were also sampled for mosquitoes were tested for *Plasmodium* species. In RIMDAMAL II, serially collected DBS were collected from participants of all ages from households that were also sampled for mosquitoes, and DBS from 100 of these randomly chosen participants were tested for *Plasmodium* species. DNA was extracted from 3mm hole punches of each DBS by incubating them overnight in a 60°C shaking incubator with PBS and proteinase K and the supernatant process with a Mag-Bind Viral DNA/RNA 96 kit (Omega Bio-Tek) using the KingFisher Flex Magnetic Particle Processor (Thermo Fisher Scientific). DNA was eluted in 30 μL of nuclease-free water and stored at −80° C prior to testing.

### DNA extraction from RIMDAMAL II mosquito tissues

Female *Anopheline* head+thoraces stored in RNALater® (Thermo Fisher Scientific) were removed from tubes and blotted on absorbent paper to remove excess liquid. Samples were then individually placed in new tubes with 300 μL mosquito diluent containing 20% of FBS, 100 μg/mL of streptomycin, 100 units/mL of penicillin, 50 μg/mL gentamicin, and 2.5 μg/mL fungizone in PBS. Samples were homogenized by a single zinc-plated, steel 4.5 mm bead for 1.5 minutes at 24 Hz before being centrifuged for 5 minutes at 14,000 x g at 4°C. DNA was extracted from all samples using the Mag-Bind Viral DNA/RNA 96 kit (Omega Bio-Tek) using the KingFisher Flex Magnetic Particle Processor (Thermo Fisher Scientific). DNA was eluted in 30 μL of nuclease-free water and stored at −80°C. Abdomen samples were placed in 2.0 mL tubes with one 4-5 mm steel bead, TL buffer, and proteinase K solution provided by the Mag Bind Blood and Tissue Kit HDQ 96 kit (Omega Bio-Tek). Each sample was homogenized (50 oscillation for 5 minutes), centrifuged (8,000 rpm for 30 seconds), and incubated in a 56°C dry bath overnight. The next day, homogenates were extracted using a KingFisher™ Flex Purification System (Thermo Fisher Scientific). DNA was eluted in 100 uL of nuclease-free water and yields were quantified using a Nanodrop device (ThermoFisher Scientific) to determine the success of extraction. Once quantified, samples were stored at −80°C until further analysis.

### *Anopheles* speciation

Morphological identification of wild-caught *An. funestus* and *An. gambiae* s.l. mosquitoes was done using standard taxonomic key in the field (117, 118). In a MiniAmp PlusThermocycler, *An. gambiae* s.l. were further speciated following PCR assay which amplified a region of the intergenic spacer 2 (119) to identify *An. gambiae*, *An. coluzzii* and *An. arabiensis*. To further speciate mosquitoes belonging to the *An. funestus* complex, a portion of the ITS2 gene was amplified following the method of Koekemoer et al. (120), and subsequently sequenced. The DNA sequences were aligned in Mesquite (121), and the *Anopheles* species was determined by comparing the sequences with those found in the GeneBank nucleotide database.

### *Plasmodium* detection in both human and mosquito samples

To detect the presence of *Pf, Po, Pm* and *Pv* in the head+thorax and/or abdomen tissues, two real-time PCRs were done. *Pf* DNA was detected using the assay described by Hoffman et al. (122), which detects *P. falciparum* DNA in mosquito tissues by amplifying the *var* gene acidic terminal sequence (*var*ATS). *Po, Pm* and *Pv* were detected following the protocol described by Phuong et al. (123), which targets the 18S rRNA gene. *Pf* samples that amplified at or before 36 cycles and showed a logarithmic amplification curve were interpreted as positive. The positive controls for *Pf* were DNA from *Plasmodium*-infected DBS generated from *in vitro* culture at various ranges of parasitemia placed on filter paper, from which 3 mm punches were placed in PCR tubes with 70 μL Rapid Extraction Solution (Thermo Fisher Scientific) and rocked overnight at 4°C and the DNA was purified using the same procedure as described for head+thorax. Negative controls were DNA extracts from malaria-negative DBS. *Po, Pm* and *Pv* were detected following the protocol described by Phuong et al. (123), which involved using species-specific forward primers and probes targeting the 18S rRNA gene. *Po* was identified using a probe labeled with VIC at the 5’ end and a minor groove binding quencher at the 3’ end. *Pv* was detected using a probe labeled with TAMRA at the 5’ end and a BHQ-2 quencher at the 3’ end. *Pm* was identified using a probe labeled with FAM at the 5’ end and a minor groove binding quencher at the 3’ end. The same reverse primer was used for all species identification. Nuclease-free water served as the negative control, while linearized, clean DNA from the plasmid clones (MRA numbers 177–180; BEI Resources, NIH, NIAID) acted as both positive controls. All head+thorax samples that showed logarithmic amplification at or before 36 cycles were interpreted as positive. All PCR reactions were performed in a QuantStudio 3 real-time PCR instrument (Thermo Fisher Scientific).

### Blood meal analysis

A conventional PCR followed by a 1.5% electrophoresis gel was done to analyze the host blood meal of mosquito abdomen samples, following the methods described by Kent et al. (124), which amplifies a region of the cytochrome b mitochondrial gene and includes primers to identify blood meals from dog, human, cow, pig, and goat host species. The electrophoresis gel ran at 90 volts for 45 minutes and was imaged on a BIO-RAD ChemiDoc MP Imaging System for analysis.

### Statistical analysis

Data was analyzed on GraphPad Prism or R (R-project.org). Chi-square tests and Fisher’s exact test were performed, depending on the number of variables. Chi-square test for trend tests for linear trends between variables, while the Fisher’s exact test tests for association between variables, and both assess the chances of observing the trend/association in random sampling (GraphPad Prism Version 9.3.1). A P value less than 0.05 was interpreted as statistically significant.

## Data Availability

All RIMDAMAL II data will be deposited in ClinEpiDB.org and be available for analysis and download under the terms of set by the website.

## Notes

### Competing Interest Statement

The authors have declared no competing interest.

### Clinical Trial

NCT02509481 & NCT03967054

### Clinical Protocols

https://clinicaltrials.gov/study/NCT02509481?term=RIMDAMAL&rank=2

https://clinicaltrials.gov/study/NCT03967054?term=RIMDAMAL&rank=1

### Funding Statement

The RIMDAMAL study was funded by grant OPP1116536 from the Bill and Melinda Gates Foundation and the RIMDAMAL II study was funded by grant U01AI138910 from the U.S. National Institutes of Health

### Author Declarations

Ethics committee/IRB of Colorado State University gave ethical approval for this work (IRB numbers: 15-5796H & 18-8083H) Ethics committee/IRB of Comite d'Ethique Institutionnel pour la Recherche en Sciences de la Santé gave ethical approval for this work (EC numbers: A03-2015/CEIRES & A031-2018/CEIRES)

